# Remodelling of the gut virome after long-term fasting

**DOI:** 10.64898/2025.12.19.25342476

**Authors:** Natalie Falshaw, Quinten R Ducarmon, Alexandra King, Franziska Grundler, Robin Mesnage

## Abstract

**Background:** Long-term fasting is a promising strategy for improving human health and reducing cardiometabolic risk. Emerging evidence suggests that the gut microbiome may mediate many of these benefits, but the role of its viral component, dominated by bacteriophages, remains poorly understood.

**Methods:** Using shotgun metagenomic data from a single-arm, monocentric fasting intervention, this study profiled the gut virome (n=89 individuals, n=241 samples) before and after 9.8 days of fasting (∼250 kcal/day) as well as one and three months afterwards to examine viral population dynamics and their role in microbiome restructuring and host health.

**Results:** Fasting induced a transient loss of viral diversity and a shift toward virulent lifestyles consistent with prophage induction. External dataset validation identified 49 phages that were reproducibly differentially abundant at the end of the fast. Many were linked to bacterial hosts, showing concordant shifts, including depletion of *Faecalibacterium*-associated phages and enrichment of *Bacteroides*-associated phages. Cross-domain network analyses revealed denser, more cohesive viral-bacterial networks at the end of fast, with enriched connections to key butyrate producers, such as *Faecalibacterium* and *Roseburia* species, suggesting phages reinforce health-associated taxa as ecological anchors during fasting-induced restructuring. Fasting-induced microbiome restructuring is distinct from inflammatory disease-associated patterns.

**Conclusion:** Collectively, these findings indicate that fasting remodels cross-domain associations through reproducible, functionally relevant phage-host interactions, with reorganisation persisting for up to three months and aligning with improvements in cardiometabolic health. More broadly, these findings position the gut virome as a dynamic and structured component of dietary responsiveness, hinting that phage-host systems may play an underappreciated role in coordinating adaptive microbiome responses to metabolic interventions.

## Introduction

Fasting, whether practiced intentionally for cultural, religious, or therapeutic purposes, or encountered involuntarily through food scarcity and seasonal cycles, is a deeply conserved human experience^1^. Long-term fasting, defined as voluntary abstinence from food for more than four consecutive days^2^ typically spanning 4-21 days^3^, triggers co-ordinated physiological, metabolic^4^, biochemical^5^, and immunological adaptations^6,7^. Mounting evidence suggests these adaptations may contribute to improved cardiometabolic health^6^, healthspan^8^, and possibly lifespan^9,10^.

The gut microbiome (GM) describes the diverse community of microorganisms inhabiting the colon, their collective genome, and combined metabolic activity^11^. Diet is a primary determinant of its makeup, providing substrates for bacterial growth^12^. Accordingly, fasting induces a rapid compositional shift whereby bacteria reliant on host-derived substrates (i.e. mucins) proliferate, whilst those dependent on exogenous dietary inputs (i.e. dietary fibre) decline^13,14^. As a complex system shaped by competitive and cooperative interactions, the GM undergoes ecosystem-wide shifts in composition and function during significant caloric restriction^14,15^. This occurs in parallel with host metabolic adaptations^16^. Previous research has implicated the GM as a possible mediator of fasting-induced benefits^13^, including improved insulin sensitivity^16^ and immune regulation^17^. However, the durability of these effects likely depends on food reintroduction, as reduced microbial density during fasting opens ecological niches that recolonisation can either reinforce or destabilise^18,19^.

One principal mechanism underpinning host-microbiome interactions is microbial metabolism whereby bacteria-derived bioactive compounds, or “post-biotics”, act as signalling molecules, with systemic effects^20^. Well-characterised examples include short chain fatty acids (SCFAs) such as butyrate, produced by dietary fibre fermentation, which support gut barrier integrity and immune tolerance^21,22^, and indole-3-propionic acid (IPA), a tryptophan-derived metabolite with antioxidant and anti-inflammatory properties relevant to cardiometabolic health^23,24^. Thus, the metabolome serves as a powerful readout to connect microbial shifts with health related functional outcomes in the host^25^.

Most microbiome research to date has focused on bacteria^26^, thereby neglecting other intestinal inhabitants, such as viruses^27,28^. The gut virome is largely composed of bacteriophages (phages), which are viruses infecting bacteria rather than human cells^29^. Phages are recognised as fundamental regulators of GM structure and function^30^; by modulating bacterial populations, transferring genes, and shaping microbial metabolic outputs, phages influence both microbial ecology and, indirectly, host physiology^31^. Phages exhibit two principal lifestyles: virulent phages hijack bacterial machinery to replicate and lyse their hosts, directly reshaping bacterial populations, meanwhile temperate phages integrate into bacterial host genomes as dormant prophages in a lysogenic state^32^. Here, temperate phages can transfer genes that expand bacterial metabolic repertoires^33,34^, confer antibiotic resistance^35^, or modulate virulence^36^. This makes phages a genetic reservoir, equipping bacteria with traits that can be either beneficial^37^ (e.g. immune stimulation, metabolic resilience) or detrimental^38^ (e.g. toxin production) to human health. Prophages can later be induced, re-entering the lytic cycle in response to certain triggers^39^. Collectively, these dynamics highlight the phageome as a central but underexplored layer of host-microbiome interactions^40^.

Characterising the gut virome remains technically challenging. Viruses lack universal genetic markers, display high sequence diversity, and evolve rapidly, leaving much of the viral sequence space as biological “dark matter”^41^. Only a few thousand viruses have been isolated, sequenced, and characterised, further limiting the ability to identify and quantify viruses in the GM^42^. Two main strategies are used to sequence the virome: (1) virus-like particle (VLP) enrichment, a wet-lab technique which increases the proportion of viral sequences recovered, but this can introduce methodological biases^43^; and (2) bulk metagenomic sequencing and subsequent computational separation of viral reads, avoiding enrichment bias but often producing uneven coverage and underrepresentation of certain viruses^44,45^. To profile the virome, a reference-based approach may be used, which maps reads to viral genome databases, enabling efficient identification of known viruses, but misses novel or poorly catalogued sequences. Alternatively, an assembly-based approach involves reconstructing viral genomes *de novo* from sequencing reads, facilitating the discovery of new viral genomes, but is computationally demanding and sensitive to assembly quality. The combination of sequencing strategy and computational approach affects which viral populations are detected and how they are quantified. This is important to consider when contextualising findings withing the broader field^46,47^ and partially explains why the gut virome remains incompletely characterised.

In this study, bulk gut metagenomic data, previously analysed from a bacteriome perspective^14^, and complemented by two external validation fasting cohorts, were re-examined to investigate virome composition and dynamics at baseline, at the end of the fast, and during the 1- and 3-month follow-up periods after long-term fasting. Focusing on the phageome, while situating it within the broader microbial ecosystem^48,49^, the aim was to determine how viral populations respond to long-term fasting and assess their potential role in shaping microbiome structure, cross-domain interactions, and fasting-induced host benefits.

## Methods

### Study design and data source

This analysis utilised publicly available metagenomic sequencing data from two fasting studies at the Buchinger Wilhelmi Clinic, Germany: the OralFast observational trial^50^ and the GENESIS interventional study^51^. Full study protocols have been described previously^14^. Briefly, 89 participants (Table S1) underwent a medically supervised fasting programme lasting an average of 9.8 days at 200-250 kcal/day, followed by 3-4 days of structured food reintroduction. A subset from GENESIS provided follow-up data at one (n=32) and three months (n=31; Fig.1). Anthropometric and clinical measures were taken daily. Venous blood was collected at baseline and the end of fasting. Faecal samples were self-collected and frozen within 12 hours, except for follow-up samples which were returned by post, and all samples underwent shotgun metagenomic sequencing on the Illumina NovaSeq 6000 platform. Paired-end FASTQ files used for downstream bioinformatic analysis were quality filtered, and host decontaminated (Fig.S1A), and 241 gut metagenomes (371 GB) were retained for taxonomic profiling.

**Figure 1:**
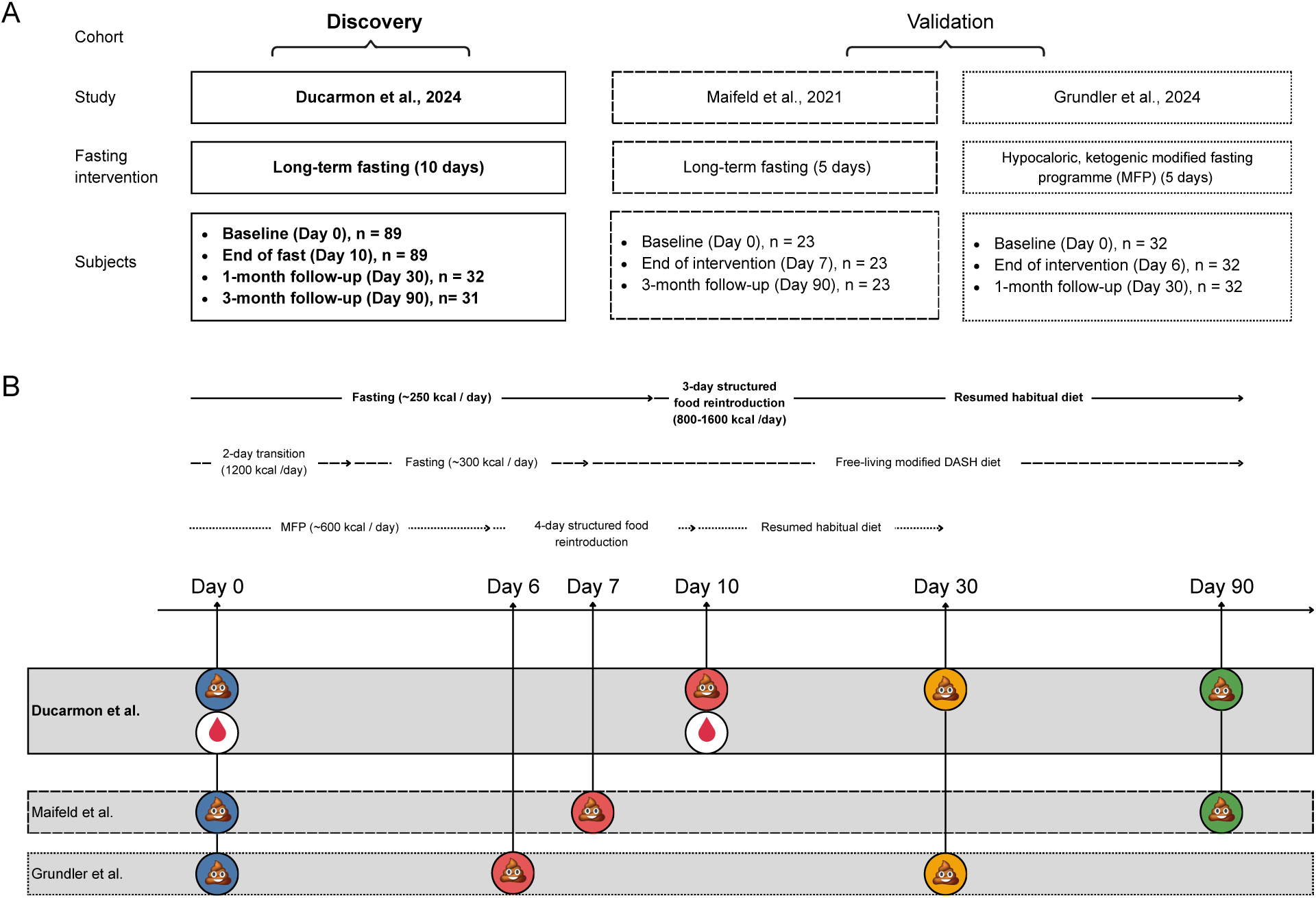
Study design and sample collection timeline for discovery and validation cohorts. (A) Overview of included studies. The discovery dataset comprised participants from clinical trials conducted at the Buchinger Wilhelmi Clinic, Germany (Ducarmon et al., 2024). Participants underwent a medically supervised long-term fasting programme (∼10 days; 200-250 kcal/day) with structured refeeding. Stool samples were collected at baseline (Day 0), end of fast (Day 10), and at follow-up (1 month, n=32; 3 months, n=31). Validation datasets were also re-processed using the same pipeline. Maifeld et al. (2021) studied patients with metabolic syndrome (n=23) undergoing a 5-day fast followed by a modified Dietary Approach to Stop Hypertension (DASH) diet, with samples collected at Day 0, Day 7, and Day 90. Grundler et al. (2024) included healthy individuals (n=32) completing a 5-day hypocaloric ketogenic modified fasting programme (MFP) at home, with stool samples at Day 0, Day 6, and Day 30. (B) Schematic timeline of interventions and sampling points across the three cohorts. Icons indicate stool (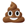) and blood ( 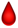 ) collections. Study stages are labelled along horizontal arrows with dashed borders, where the pattern to the cohort represented.

### External validation cohorts

For validation and replication, two independent longitudinal metagenomic datasets were included^52,53^ (Fig.1). Sequencing protocols in the validation cohorts were either identical^53^ or highly comparable^52^.

### Taxonomic profiling

Bioinformatic processing was conducted via CLIMB^54^ (Cloud Infrastructure for Microbial Bioinformatics), a UK-based cloud-based high-performance computing platform designed for microbial genomics. FASTQ files were processed using Phanta (v1.1.0) a non-assembly k-mer based bioinformatic pipeline optimised for viral and microbial profiling against a comprehensive, gut-specific reference database^55^ (Fig.S1B). Default parameters were used including a Kraken2 classification confidence threshold of 0.1 to reduce ambiguous assignments. Reads were assigned to the lowest possible taxonomic level, followed by estimation of genome coverage to exclude low-confidence hits. Taxa were only retained if coverage exceeded 10% for viruses and 1% for bacteria. Classification statistics summarising read counts, classification efficiency, and number of reads assigned were produced during Phanta execution and used to confirm adequate sequencing coverage across all samples. With the exception of alpha diversity metrics, which used raw counts, relative taxonomic abundance tables formed the input for all downstream analysis. Both the discovery and validation cohorts were processed using this same bioinformatic pipeline to minimise effects of bioinformatic processing.

### Post-processing: lifestyle, viral-host mapping and cross-domain correlation analysis

Viral lifestyles (temperate versus virulent) were predicted using BACterioPHage LIfestyle Predictor (BACPHLIP)-based annotations, as implemented in Phanta’s post-processing pipeline^55,56^. Phanta’s host-collapsed output, which links viral species to a predicted bacterial host genus using integrated Phage-Host Prediction (iPHoP) assignments, was used to map phages to their inferred bacterial host genera^55,57^. Cross-domain co-abundance associations between bacterial and viral genera, independent of direct phage-host pairings, were inferred using FastSpar^55,58^. Analyses were performed at the genus level to balance taxonomic resolution and interpretability. Only participants with complete longitudinal sampling across all four timepoints (n=28) were included in the network analysis. Within each timepoint, only taxa present in more than 20% of samples were retained. FastSpar was run with 500 bootstrap permutations per timepoint and resulting correlation and p-value matrices were filtered to retain only significant (*p*<0.05) cross-domain (virus-bacteria) associations (Fig.S1C).

### Statistical analysis

All statistical analyses and data visualisation were performed in R (v4.2.3). To account for the longitudinal design and repeated measures, participant identity was included as a stratification factor in PERMANOVA (β diversity) and as a random effect in all linear mixed-effects models (LMMs) (Fig.S1D). The general model structure was as follows: *Response ∼ Timepoint (1|Participant)*.

Changes in twelve clinical biomarkers, reflecting cardiometabolic, hepatic and inflammatory function, were tested between baseline (Day 0) and the end of fast (Day 10) using paired Wilcoxon signed-rank tests. *P*-values were adjusted for multiple comparisons using the Benjamini-Hochberg (BH) procedure to control the false discovery rate (FDR), and adjusted values are reported as *q*-values.

Only annotated viral taxa were retained for downstream analysis. Alpha diversity (Shannon index and observed richness) was calculated from raw count data using the *vegan*^59^ package. Distributions were visually inspected and assessed for skewness to confirm suitability for linear modelling. Differences across timepoints were evaluated using LMMs.

Relative taxonomic abundance tables were normalised to per-sample relative abundances prior to beta diversity and differential abundance analyses. Beta diversity (between-sample diversity) was determined using Bray-Curtis dissimilarity visualised by principal co-ordinates analysis (PCoA) and tested by permutational multivariate analysis of variance (PERMANOVA) with pairwise comparisons adjusted for multiple testing with BH. To assess potential confounding or effect modification, interaction models were run between timepoint and participant covariates, including sex, age, fasting duration, and BMI. Virulent and temperate community lifestyle summaries were compared with LMMs. Differential abundance testing was performed with *MaAsLin2*^60^, using per-sample viral relative abundances as inputs and applying log transformation within the model. Timepoint was included as the fixed effect (baseline as reference). Multiple testing correction was applied using BH FDR, and viral species were considered significant at *q*<0.05.

### Viral-host directionality analysis

For the bacterial component of the discovery cohort, published mOTUs-based analyses of the same metagenomic data^14^ were re-aggregated to the genus level, transformed to relative abundances, and re-analysed with *MaAsLin2* to enable direct comparison with phage abundance changes relative to their predicted bacterial host. Viral and bacterial changes were then integrated at the level of the predicted host genus, with manual harmonisation of genus labels where required, to assess directionality of fasting-induced phage-host dynamics.

### Cross-domain network analysis

FastSpar correlation matrices were converted into edge lists and filtered at an absolute Spearman’s rho (|ρ|>0.35) to prioritise biologically meaningful interactions. Each edge represents a significant correlation between viral and bacterial genera, with positive edges indicating co-occurrence, and negative ones suggesting exclusion or predator-prey-like dynamics. Networks were classified as acute, delayed, or persistent based on based on presence-absence patterns across timepoints. For visualisation, up to the 250 strongest edges per timepoint were plotted using a shared force-directed layout to enable direct comparison of network architecture across conditions. Nodes represented taxa, and hubs were highly connected nodes central to the network.

## Results

### Subject characteristics

The cohort consisted of 89 mixed-sex, generally healthy, middle-aged participants with an average BMI in the overweight range. All participants underwent a medically supervised fasting programme lasting on average 9.8 days with ±250 kcal / day (Table 1).

**Table 1:**
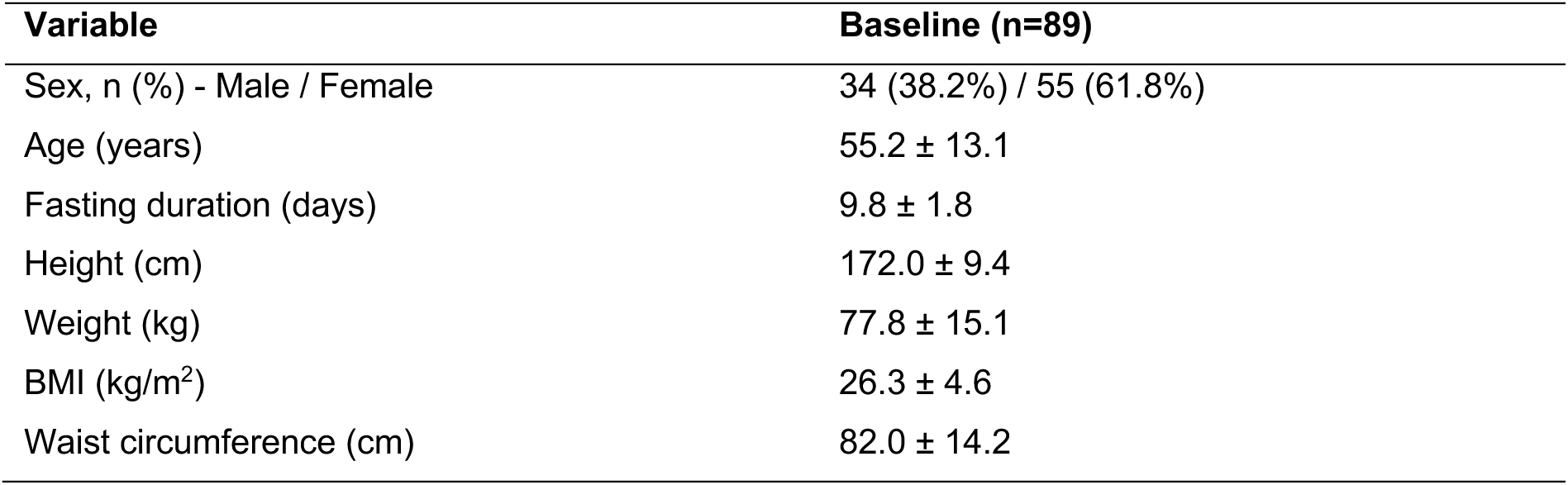
Participant characteristics at baseline prior to the fasting intervention. Values are presented as mean (± standard deviation) unless otherwise stated.

### Clinical impacts of fasting

As originally presented for this cohort, fasting induced significant shifts across markers of glycaemic control (glucose, insulin, and HbA1c) and lipid metabolism (triglycerides, HDL, and LDL), associated with improvements in cardiometabolic health (Table S2 Fig.S2; *q*<0.05). Body weight dropped by 5.02 ± 1.60 kg, and waist circumference declined by 5.59 ± 3.57 cm indicating preferential loss of visceral fat^61^. Markers of liver function (e.g. gamma-glutamyl transferase) improved^62^, decreasing by 5.33 ± 8.45 U/L, while renal indices showed expected alterations in nitrogen and purine metabolism (urea decreased by -11.14 ± 5.64 mg/dL , uric acid increased by 3.18 ± 1.98 mg/dL)^63,64^. C-reactive protein increased by 1.16 ± 6.52 mg/L, possibly reflecting acute systemic adaptation to fasting^65^, although baseline inflammation is a known determinant of CRP variability and was not stratified in these analyses^66^. These changes point to a metabolic switch from dietary substrates to endogenous energy stores, positioning fasting as systemic metabolic reset.

### Gut microbiome sequencing

Among the 241 metagenomic samples processed with Phanta, sequencing depth ranged from 6.5 to 27 million total reads per sample (median: 12.5 million). The proportion of all reads successfully taxonomically classified after quality control at the final step of the Phanta pipeline was 1.29-31.02%, median: 3.71%. Of this classified fraction, 6.4-98.0% (median: 93.6%) were viral , reflecting Phanta’s comprehensive database of gut viral sequences. No samples were excluded on the basis of read depth or classification efficiency.

Across all metagenomes, 10,863 viral species were detected, with prokaryotic viruses dominating the virome (99.5%), consistent with previous reports^47,67^. As in other large-scale gut virome studies^68–70^, crAss-like viruses were prevalent, found in 78.8% of samples, with a median abundance of 0.8% but reaching up to nearly 40% of the virome in some individuals. Taxonomically, 57% (6190) viral taxa could be assigned to a known viral family, whilst the remaining 43% (4673) were unclassified at the family level. In line with previous research, the most prevalent annotated viral families across all samples were *Siphoviridae* (76.3%), *Myoviridae* (12.5%), *Podoviridae* (7.8%) and *Microviridae* (2.1%)^47,67,71^.

### Gut virome diversity and restructuring after fasting

Both markers of alpha diversity significantly decreased at the end of the fast (Day 10; Shannon index: β=-0.057, *p*<0.001; richness: β=-38.8, *p*<0.001). However, follow-up timepoints were not significantly different from baseline (Fig.2A+B), indicating a transient reduction in virome complexity after fasting, which seems to partially recover over time. Principal co-ordinates analysis (PCoA) of Bray-Curtis dissimilarities showed PCoA1 and PCoA2 explained 4.6% and 3.8% of the variance, respectively. Visual inspection of the PCoA plot suggested a shift in virome composition after the fasting intervention, particularly between baseline and end of fast (Fig.2C). PERMANOVA confirmed a statistically significant effect of fasting on the overall virome structure (R^2^=0.02, *p*=0.001). Pairwise PERMANOVA tests revealed the largest and most statistically significant shift was observed between baseline and end of fasting (R^2^=0.01, *q*=0.002). Significant differences were also detected between end of fast (Day 10) samples and both 1-month follow-up (R^2^=0.02, q=0.002) and 3-month follow-up (R^2^=0.01, q=0.002). In contrast, no statistically significant differences were observed between before and either of the follow-up timepoints, nor between 1-month and 3-month follow-up. To ensure these effects were not driven by participant-level covariates, interaction models were tested for sex, age, fasting duration, and BMI, none of which showed significant effects or interactions with timepoint. This supports that the observed compositional changes were attributed to fasting, rather than confounded by demographic or physiological variation, and that virome restructuring was most pronounced at the end of the fast (Day 10), with partial restoration during follow-up.

**Figure 2:**
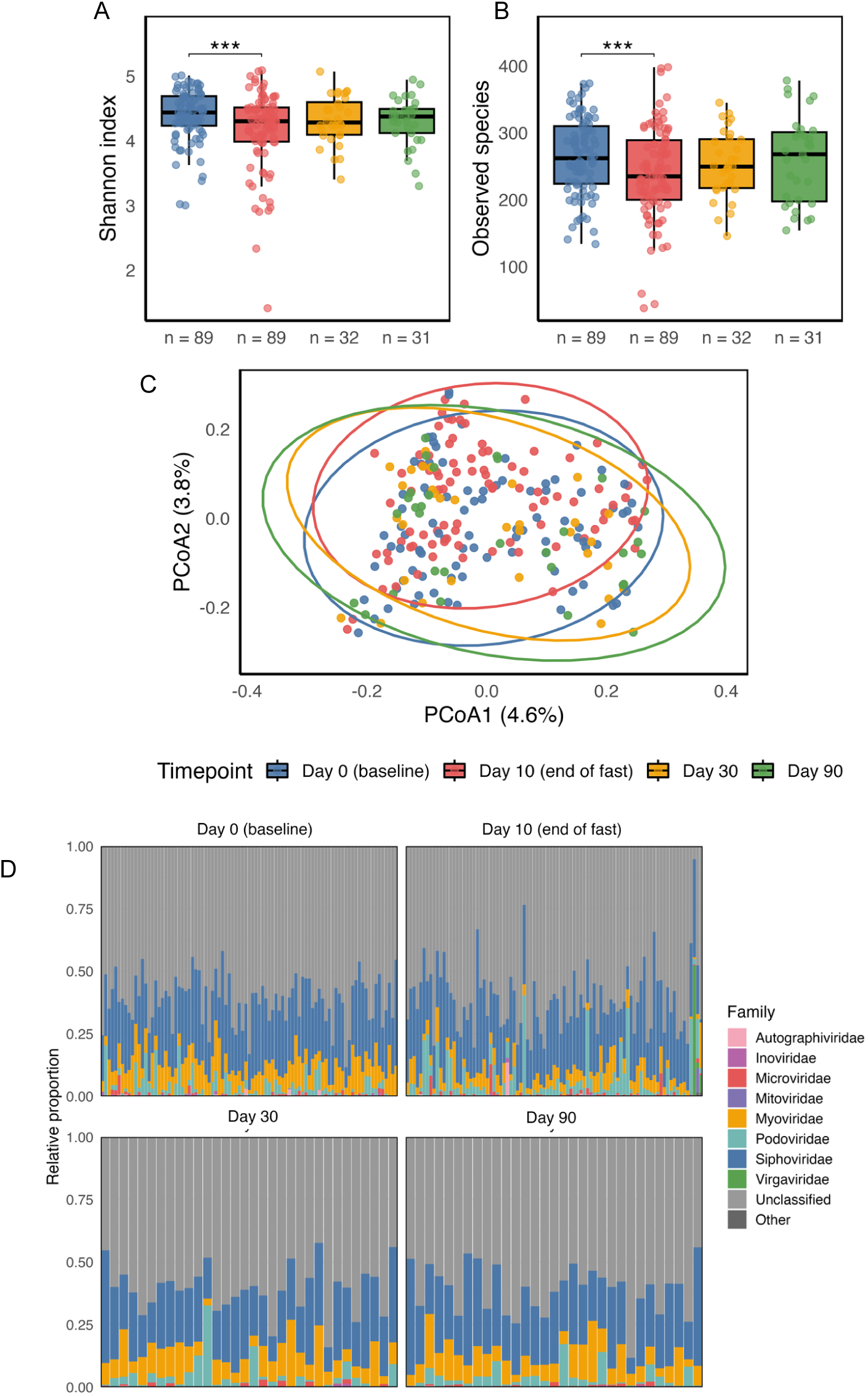
Fasting-induced changes in gut virome diversity and community structure. Alpha diversity across timepoints, show as (A) Shannon index and (B) observed species richness across timepoints, with significance assessed using linear mixed-effects models (\*\*\**p*<0.001). (C) Beta diversity visualised by Principal co-ordinates analysis (PCoA) of Bray-Curtis dissimilarities. Each point represents an individual sample, and coloured ellipses indicate 95% confidence intervals. PERMANOVA was applied to test for differences in community composition across timepoints. (D) Family-level composition of the gut virome across timepoints. Stacked bar plots show the relative abundance of viral families within each sample (Day 0 and Day 10: n=89; Day 30: n=32; Day 90: n=31).

Visualisation of relative viral family abundances over time highlight consistent dominance of *Siphoviridae* and *Myoviridae,* with individual responses and fluctuations at the end of the fast but with no clear family-level enrichment or depletion over time (Fig.2D).

Across all paired samples, fasting also induced clear shifts in phage lifestyle balance (Table S3; Fig.3). The virulent-to-temperate ratio increased significantly at the end of the fast (Day 10), both when measured by species counts (Fig.3A) and by relative abundance (Fig.3B). Assessing richness provided further resolution on what drove this shift; virulent phage richness did not change significantly across timepoints (Fig.3C). In contrast, temperate phage richness declined by the end of the fast (Day 10) and remained reduced at 1-month follow-up, before returning towards baseline by 3-month follow-up (Fig.3D). This reduction in temperate phage diversity, rather than any change in virulent richness, accounts for the altered virulent to template ratio. Compositional patterns were consistent with these richness treads. Virulent phages increased significantly in relative abundance at the end of the fast (Fig.3E), whereas temperate phage relative abundance showed comparatively little change across timepoints (Fig.3F). Together, these findings indicate that fasting shifts the gut virome towards a more virulent lifestyle signature, characterised by reduced temperate diversity and increased relative representation of virulent phages.

**Figure 3:**
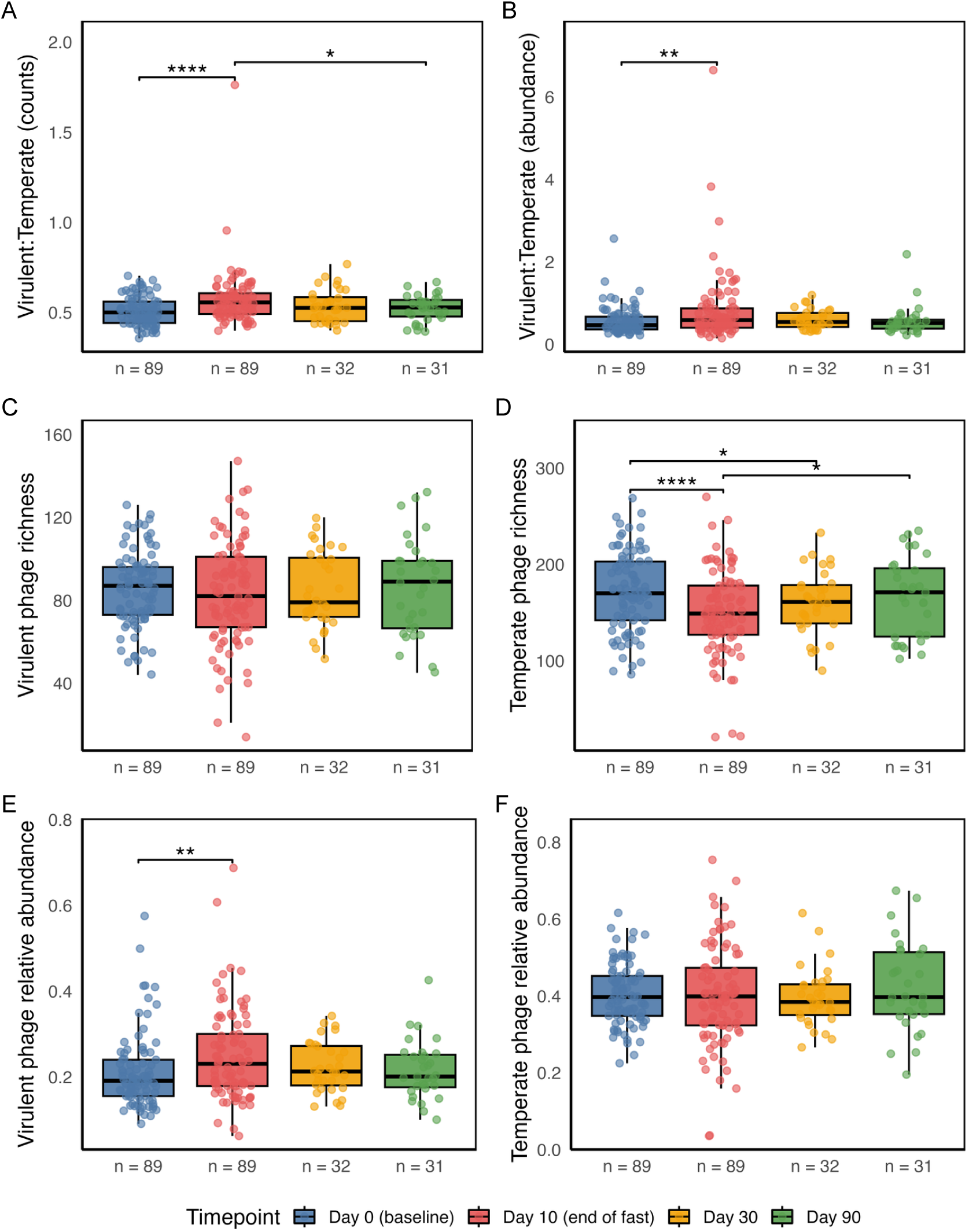
Viral lifestyle dynamics across fasting and follow-up. Boxplots show the ratio of virulent to temperate phages (top row), richness (middle row), and relative abundance (bottom row) at baseline, end of fast, and follow-ups. Each point represents an individual sample. Sample sizes per timepoint are indicated below plots. Significance was assessed using linear mixes-effects models with FDR correction ( \**p*< 0.05; \*\**p*< 0.01; \*\*\*\**p*<0.0001).

### Fasting-associated viral signatures

Of 10,863 viral taxa, 121 were differentially abundant (*q*<0.05), all with a prevalence of at least 10% (Table S2) between baseline (Day 0) and the end of the fast (Day 10) (Table S4). The 25 most significantly altered taxa are shown by model coefficients (Fig.4A) and their distribution across individuals and timepoints in a heatmap (Fig.S3). Despite inter-individual variability, consistent enrichment and depletion patterns were evident, with partial but non-uniform recovery towards baseline at later timepoints. Collapsing all 121 significant viruses to families revealed divergent, within-family responses (Fig.4B): both increases and decreases occurred among *Siphoviridae*, *Myoviridae*, and unclassified viruses, with a single *Microviridae* feature decreasing. Therefore, fasting seems to non-uniformly shift families and redistribute constituent species in opposite directions.

**Figure 4:**
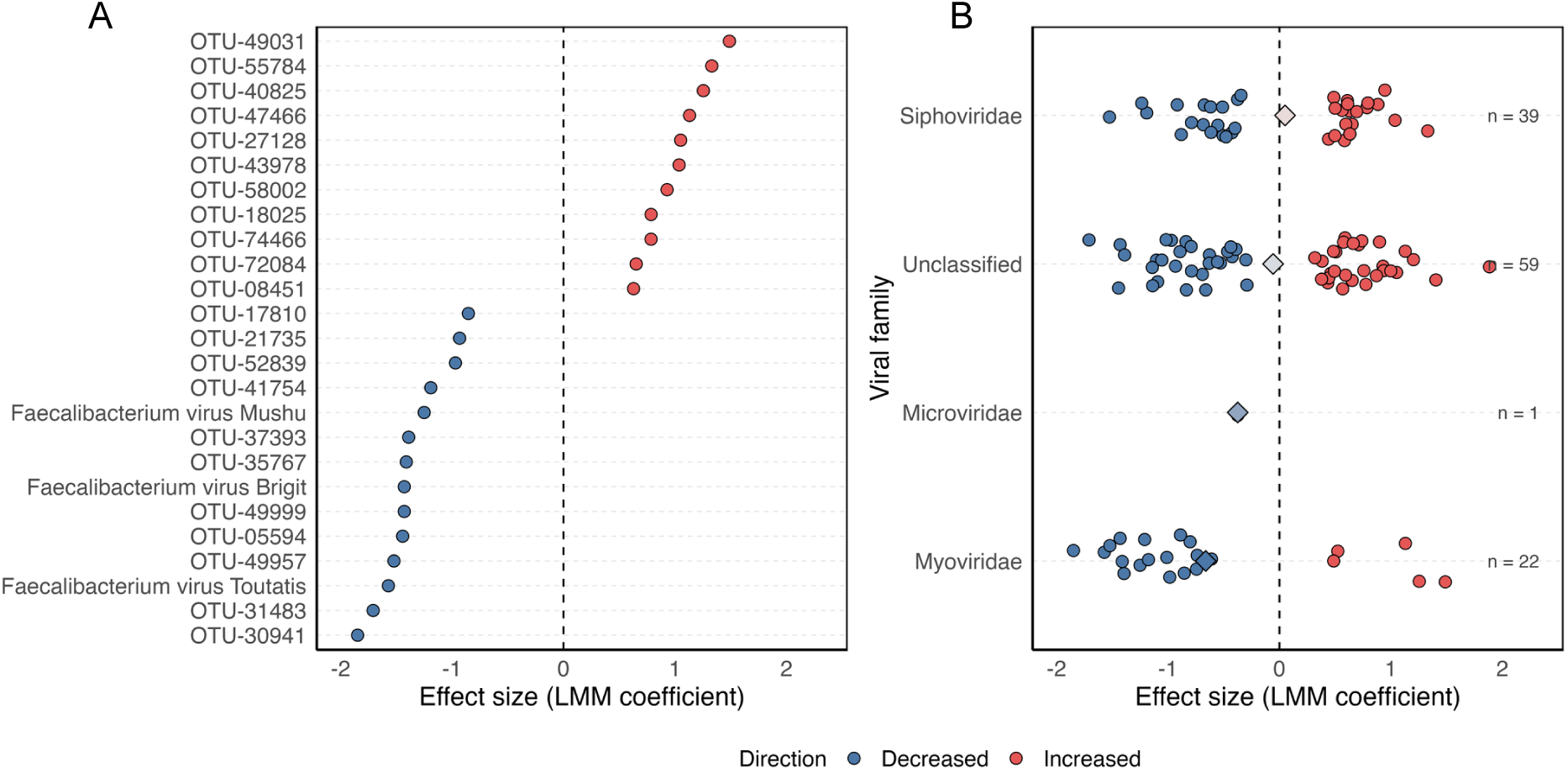
Fasting-associated changes in viral taxa. (A) Top 25 viral species significantly associated with fasting, identified using linear mixed-effects models. Species are ordered by model coefficient (log fold change in relative abundance at end of fast vs. baseline), with points coloured by direction of change. (B) Family-level responses to fasting, showing all 121 significant species collapsed to viral families. Individual species are plotted as points, with diamonds indicating mean family-level coefficient. Labels indicate the number of significant species per family.

### Validation

The present study showed the strongest post-intervention restructuring, with Bray-Curtis dissimilarity reflecting whole-community compositional change peaking at the end of the fast (Day 10), declining by 1-month follow-up, and rising slightly by 3-month follow-up, consistent with bacterial trends in the same cohort^14^ (Fig.5A). In contrast, Maifeld et al. showed minimal divergence, and Grundler et al. displayed an intermediate trajectory, with moderate changes at the end of the fast (Day 7) followed by recovery.

**Figure 5:**
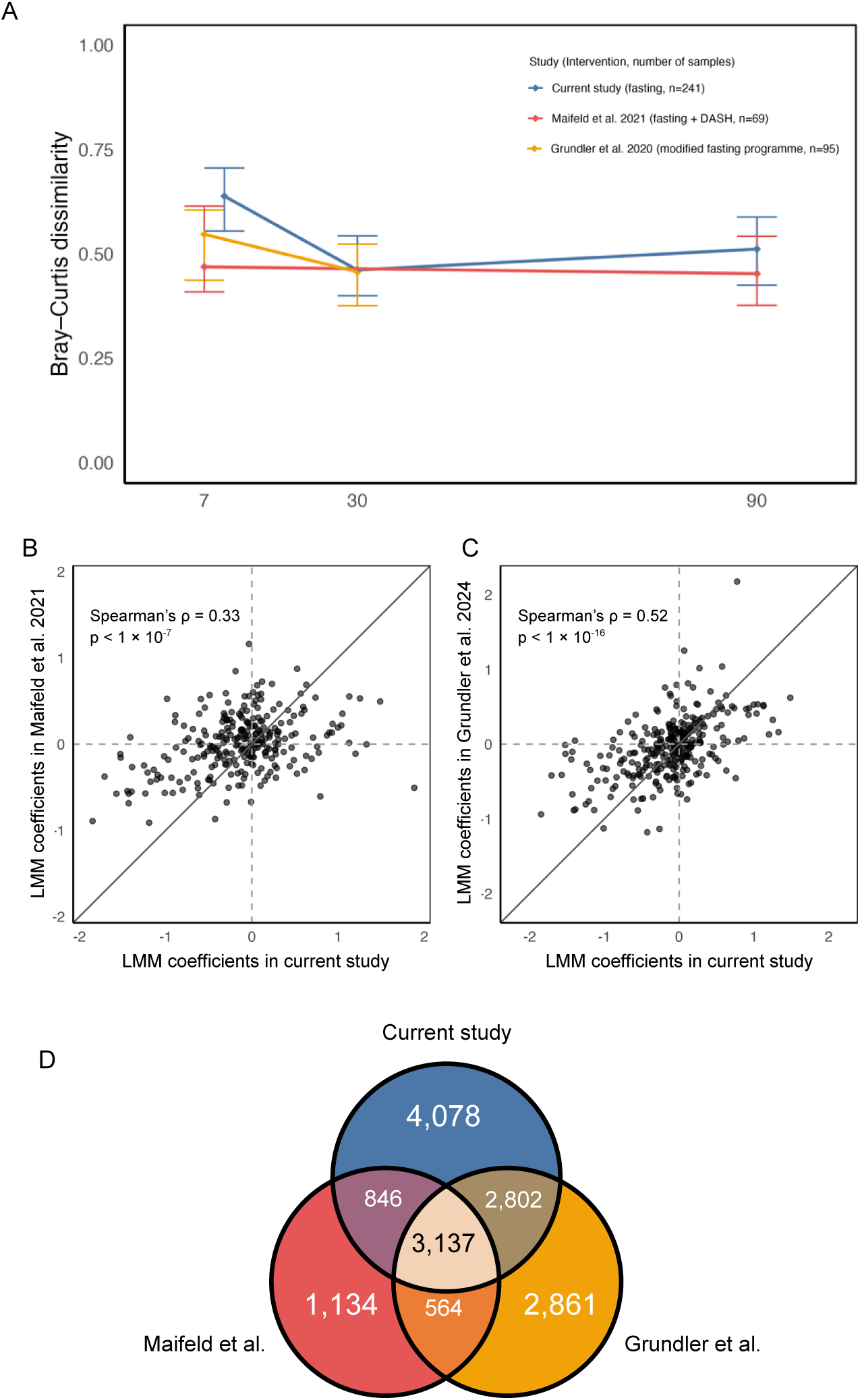
Cross-cohort validation of fasting-induced virome shifts. (A) Longitudinal fluctuations in Bray-Curtis dissimilarities comparing end of fast and follow-up samples to baseline in discovery and validation cohorts. Values near 0 indicate similarity to baseline, while values approaching 1 reflect divergence. Lines show median dissimilarity with interquartile range. (B) Replication of viral differential abundances between the discovery cohort and Maifeld et al. (2021). Linear mixed-model effect sizes are plotted for viral features detected in both studies; solid diagonal indicates perfect agreement (x = y), dashed lines mark zero effect. Spearman’s rho (ρ) and *p*-values shown. (C) Replication of viral differential abundances between the discovery cohort and Grundler et al. (2024), as in panel B. (D) Venn diagram showing shared and unique viral taxa across the three cohorts.

Cross-study comparisons demonstrated reproducibility of specific virome shifts; scatterplots of model coefficients revealed significant correlations between the discovery and validation cohorts, with stronger concordance observed with Grundler (Spearman’s rho [ρ]=0.52, *p*<1×10⁻¹⁶; Fig.5C) than Maifeld (ρ=0.33, *p*<1×10⁻⁷; Fig.5B). Quantification of shared viral features revealed both a unanimous core and substantial study-specific diversity. A total of 3,137 taxa were common to all three cohorts; 2,802 were shared between the discovery and Grundler, and 846 between the discovery and Maifeld, while 564 were shared by Maifeld and Grundler. Each cohort also contained unique taxa (Ducarmon: 4,078; Maifeld: 1,134; Grundler: 2,861), reflecting differences in host population, sampling, and sequencing depth (Fig.5D). However, in both validation cohorts no viral taxa reached significance following LMMs, possibly due to reduced cohort size.

In total, 49 “consensus” viral species were identified (Table S5), defined as those among the 121 significantly differentially abundant species in the discovery cohort that were also detected in both validation cohorts and consistently shifted in the same direction at the end of the fast (Fig.6A). Across the combined dataset, prevalence of consensus taxa was moderate overall (median=30.7%; IQR=17.8-44.3%), ranging from relatively rare (12.0%) to widespread (87.6%), highlighting their potential relevance as fasting-associated virome signatures.

**Figure 6:**
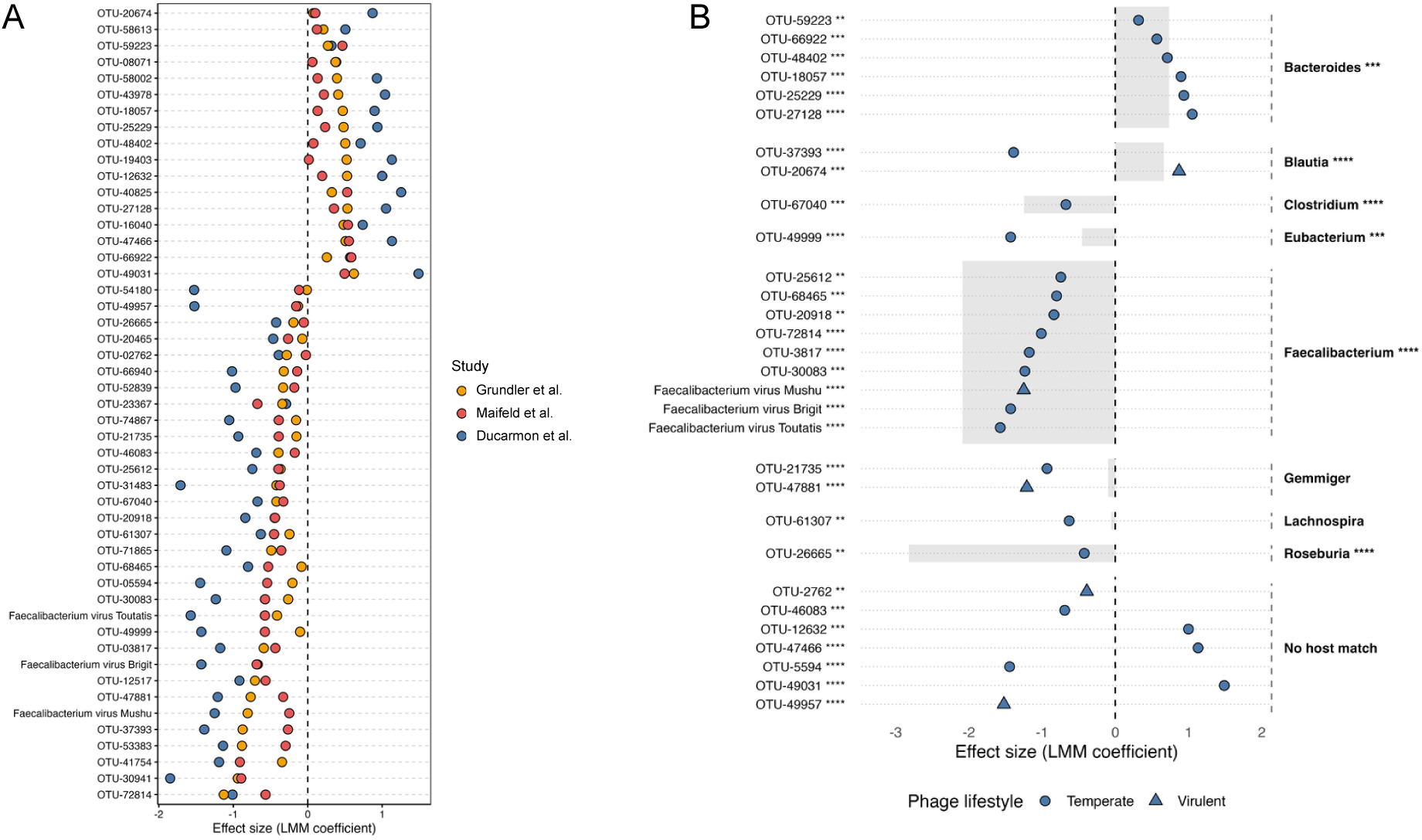
Cross-study and host-associated responses of consensus fasting-responsive viruses. (A) Cross-study comparison of top consensus viral species with consistent response to fasting. Each point represents the effect size (LMM coefficient) for a viral species’ differential abundance between baseline and end of fasting samples within one of three independent fasting cohorts. Vertical dashed line indicates no change. Horizontal grey dashed lines separate individual species. Viral species are ordered by the magnitude of their average effect size across studies, with consistently decreasing species shown at the top and increasing species at the bottom. Consensus species were defined as those significantly changing in the primary study (*q*<0.05) and detected across all three cohorts, enabling directional and cross-study consistency to be visualised. (B) Fasting-responsive viruses grouped by their predicted bacterial host genus. Points indicate viral LMM coefficients at end of fast compared to baseline, with shape denoting lifestyle prediction. Background grey bars show the corresponding bacterial host genus LMM coefficient from the same cohort and samples. Left-side labels list viral OTUs/species with FDR significance thresholds indicated (* < 0.05, ** < 0.01, *** < 0.001, **** < 0.0001). Right-side labels indicate the matched bacterial host genus, with significance shown according to bacterial FDR thresholds. Viruses without a matched genus in the complementary bacterial dataset are grouped under “No host match.”

Of the consensus fasting-responsive species, 30 could be confidently mapped to predicted bacterial hosts using Phanta annotations, enabling joint assessment of viral and bacterial responses (Fig.6B and Table S6). Most phage-host pairs shifted concordantly, particularly within *Faecalibacterium* (e.g. *Mushu, Brigit, Toutatis*), *Eubacterium*, *Roseburia*, *Clostridium* (manually aliased to account for recent reclassification of *Lacrimispora*^72^), and *Bacteroides*, consistent with phage-host coupling and co-ordinated restructuring of the virome and bacteriome during fasting. A single *Blautia* phage displayed discordant dynamics relative to its host, suggesting more complex interactions. The majority of mapped viruses were predicted to be temperate phages, supporting the idea that lifestyle flexibility underpins adaptive host-virus interactions in response to environmental perturbations^73^.

### Cross-domain network analysis

To ensure longitudinal consistency, network reconstruction was restricted to the 28 participants with samples available at all four timepoints (Table S8). Fasting induced a clear restructuring of the viral-bacterial network at the genus level (Fig.7A-C). Overall network complexity exhibited a biphasic pattern, increasing from baseline to end of fast, simplifying at 1-month follow-up (Day 30), and increasing again by 3-month follow-up (Day 90). This pattern was consistently reflected across multiple network metrics, including the total number of significant cross-domain associations (Fig.7A), network density, defined as the proportion of observed edges relative to all possible viral-bacterial connections (0.048, 0.060, 0.037, 0.060) and mean degree, defined as the average number of connections per node (14.4, 18.1, 10.8, 18.0). Across all timepoints, co-abundance correlations were predominately negative (82-94% of edges), suggesting that inverse or competitive relationships largely governed viral-bacterial dynamics. Positive associations were most frequent at 1-month follow-up (17%), when total edges were the lowest, aligning with this biphasic pattern in network complexity.

**Figure 7:**
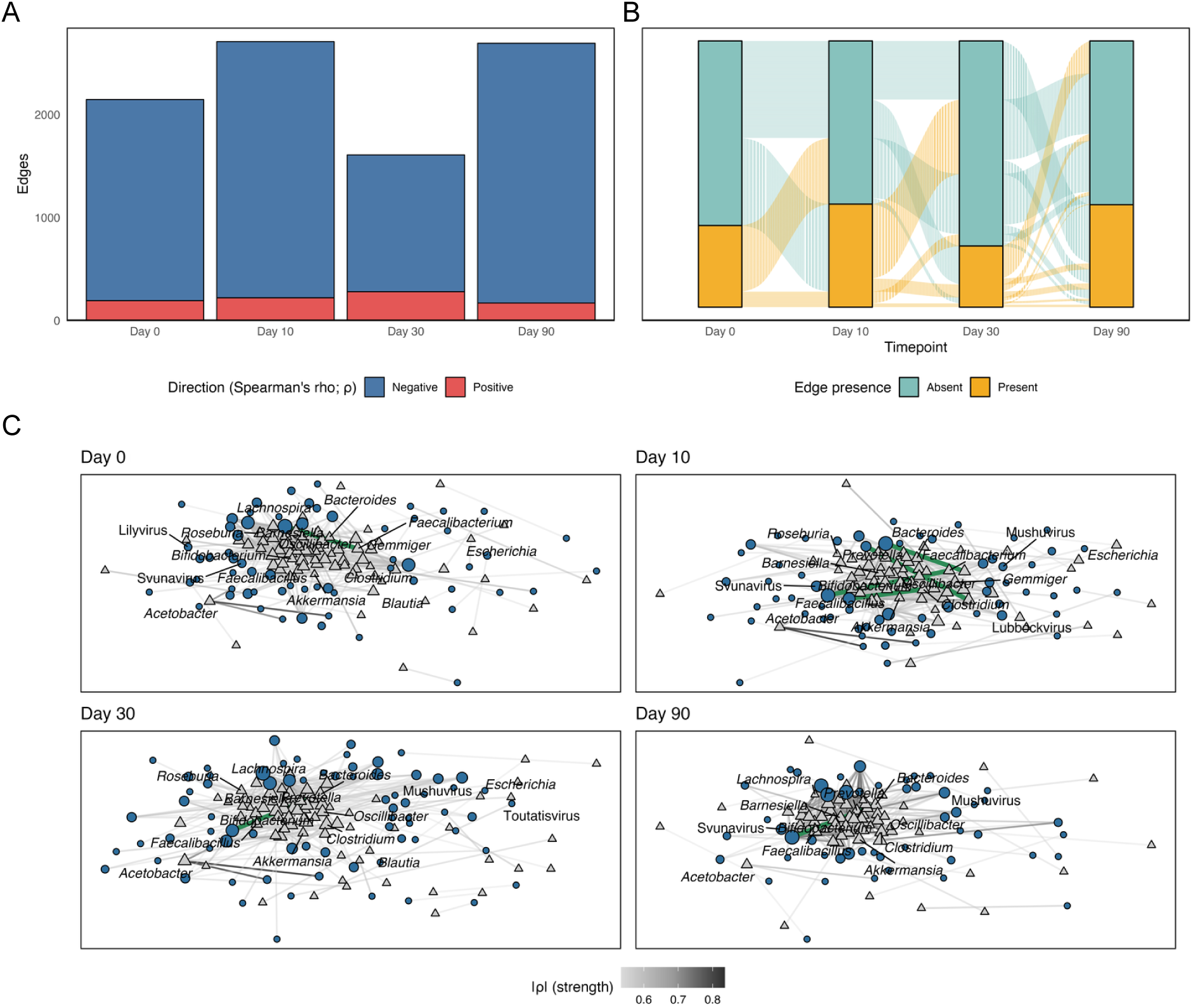
Cross-domain co-abundance network analysis across timepoints. (A) Number of significant edges by timepoint, coloured by correlation direction. (B) Edge persistence and turnover across timepoints. (C) Network layouts showing virus-bacterium co-abundance associations. Viral nodes are represented by blue circles, bacterial nodes as grey triangles. Node size is proportional to connectivity (degree), while edge thickness and shading reflect correlation strength (|ρ|); green edges highlight predicted host-phage pairs. Networks are shown for baseline (Day 0), end of fast (Day 10), and follow-up at Day 30 and Day 90. Annotated nodes indicate genera of functional interest from the bacterial analysis, consensus viral genera, or other prominent hubs.

Tracking how viral-bacterial connections appeared or disappeared over time revealed extensive temporal turnover and network plasticity (Fig. 7B). Across the 6,997 unique cross-domain edges detected, only 67 (∼1%) persisted across all timepoints. The fasting period (Day 0 to Day 10) induced the largest absolute turnover, with 2,301 novel edges emerging and 1,738 edges disappearing. This clear reconfiguration is consistent with fasting acting as a strong ecological perturbation to phage-bacterial interaction networks (Fig. 7B).

Network visualisations and node-level analyses show that fasting also alters the identity of key community taxa (Fig. 7C; Fig. S4; Table S7). While mean |ρ| remained stable across timepoints (0.44-0.45), baseline networks were comparatively sparse with few highly connected nodes, whereas end of fast and follow-up networks displayed pronounced clustering and greater interconnectivity among both viral and bacterial nodes. At end of fast (Day 10), connectivity was dominated by viral hubs such as *metagenomic gut virus-(mgv)400018*, *mgv400016*, and *Svunavirus*, each exceeding 50 edges, alongside bacterial hubs including *Solobacterium*, *Faecalibacillus*, *Akkermansia*, and *Faecalibacterium*, which reached 40-60 edges (Fig.S4). Most major hubs diminished in connectivity by the 1-month follow-up, whereas by the 3-month follow-up several bacterial taxa (*Clostridium, Streptococcus, Dialister, Oscillibacter, Bacteroides*) and viral clades (*Mushuvirus, Brigitvirus, mgv400339*) reached their highest degrees.

A small but stable subset of direct host-phage pairs (4-9 per timepoint) represented 0.2-0.3% of all edges (Fig.7C). These rare edges had comparable or stronger correlation values (mean |ρ| = 0.43-0.55) than the overall network average, indicating that verified viral-host pairs form a resilient core in the fluid cross-domain network. Cross-referencing the genus-level networks with the consensus viral dataset of 49 fasting responsive viral species showed that 31 of the 32 corresponding consensus viral genera were represented within the networks. These included *Faecalibacterium* associated phage genera such as *Mushuvirus, Toutatisvirus* and *Lilyvirus*. Although this comparison is made at a broader taxonomic resolution than the species-level consensus, the high degree of overlap indicates that the most stable constituents of the fasting-responsive virome are also those most structurally embedded within the microbial interaction network.

## Discussion

This study provides the first human investigation of the gut virome in response to long-term fasting, revealing a rapid reorganisation of virome composition and dynamics alongside viral-bacterial interactions that may contribute to the cardiometabolic benefits of fasting.

### Fasting reshapes virome composition and may induce prophages

Viral richness and Shannon diversity declined at the end of the fast but rebounded at follow-up, indicating the virome is sensitive yet resilient. The modest but significant β-diversity shift (largest between baseline (Day 0) and end of fast (Day 10), fading after) shows that fasting alters community structure without causing a complete replacement of the virome. These patterns mirror bacterial compositional changes from the same dataset^14^, and align with prior evidence that virome and bacteriome diversity track together, driven through bacteriophages rather than eukaryotic viruses^67^. In dense microbial environments, such as the gut, phages often favour lysogeny, integrating into bacterial host genomes and promoting long-term stability^74,75^. At the same time, predatory lytic phages sustain GM homeostasis^76,77^ by eliminating competing or less fit bacteria^78,79^, with downstream consequences for host immune status and metabolic health^39,80^. Prophage induction can be triggered by a range of factors^81–84^, including nutritional stress^85^. Here, the observed increase in the virulent-to-temperate ratio alongside reduced temperate diversity, suggests that fasting may destabilise lysogeny and promote prophage induction, enriching virulent phages^86–88^. Collectively, these findings indicate fasting perturbs, virome diversity, with virome and bacteriome trajectories remaining tightly coupled during ecological restructuring. The shift towards a more lytic signature may reflect transient prophage induction, though consequences for host health remain unclear.

### Consensus phage signatures and host-viral coupling

To identify reproducible viral signatures of fasting, the discovery cohort was combined with two validation cohorts to compare virome composition at baseline and end of fast (or hypocaloric, ketogenic modified fasting interventions). In total, 49 viral species shifted consistently across the discovery and validation cohorts, indicating structured, specific responses rather than stochastic fluctuations of low abundance taxa. These phages spanned a limited set of viral genera and bacterial hosts, suggesting that fasting selectively remodels specific components of the gut virome. Most phages, including all nine annotated *Faecalibacterium* phages, showed concordant abundance shifts with their bacterial hosts, consistent with phage-host coupling under lysogeny^89^. In contrast, a single *Blautia* phage displayed discordant dynamics relative to its host, which may reflect differences in induction propensity or shifts in phage lifecycle strategy under fasting-associated stress^39^. Given the well-recognised probiotic contributions of *Blautia*^90^ and *Faecalibacterium*^91^ to gut and metabolic health, the fasting-responsive shifts observed in their phages and hosts represent ecologically and clinically relevant targets for further investigation. Interestingly, prior work showed most *Faecalibacterium* prophages are enriched in inflammatory bowel disease, suggesting disease-driven prophage induction may exacerbate host bacterial depletion via lysis^92^. In contrast, fasting here was associated with a depletion in *Faecalibacterium* phages, implying phage-host dynamics under metabolic stress fundamentally differ from those observed in inflammation-associated microbial imbalances. The concurrent decline in *Faecalibacterium* abundance^14^ more likely reflects limited availability of fermentable dietary substrates^93^ rather than phage-mediated lytic destruction. This highlights how environmental context, whether driven by nutrient supply or host inflammation, modulates phage-host interactions^94^ and their downstream consequences for microbiome composition and host physiology^32^. Further, consensus phages providing reproducible virus-host links could advance the collective understanding of diet-microbiome interactions^95^. Future integration of viral, bacterial, metabolomic, and clinical data will be essential to establish whether such shifts actively influence host physiology or primarily mirror microbial reorganisation, and to determine whether phage responses to fasting and food reintroduction contribute to therapeutic modulation of the GM.

### Fasting strengthens cross-domain networks

Cross-domain co-abundance networks demonstrated clear restructuring after fasting. Edge counts increased sharply at the end of the fast, then dipped at the 1-month follow-up and surged again by the 3-month follow-up, producing a biphasic pattern in network complexity. Negative correlations dominated networks at all timepoints, a pattern consistent with resource limitation driving antagonistic or predator-prey dynamics that can positively stabilise community structure^96,97^. Furthermore, new edges continued to appear during follow-up, implying network reassembly is gradual and progressive rather than immediate. After the initial end of fast surge in connectivity, the network underwent a transient simplification by the 1-month follow-up characterised by more edges being lost than gained. By the 3-month follow-up, extensive reconnection was observed, with the cross-domain network rebounding to a strong and cohesive state. This patterns resembles observations from a 12-month time-restricted eating trial where the most successful responders developed more interactive microbial networks than less successful dieters, suggesting that the quality of reassembly may influence metabolic outcomes^98^. This contrasts with disease-associated networks in irritable bowel syndrome^99^, type 2 diabetes^71,100^, hypertension^101^, and chronic kidney disease^102^, which typically display fewer virus-bacteria links, lower node degrees, loss of connections with beneficial taxa. These data suggest that resilient GMs are characterised by denser, more integrated cross-domain networks, which fasting reinforced over time.

At the node level, some of the most connected hubs were SCFA- and IPA-producing bacteria, including *Faecalibacterium* and *Roseburia* at the end of the fast, and *Oscillibacter* during follow-up) alongside their associated phages, highlighting their role as ecological anchors within fasting-induced network reconfiguration. Several consensus fasting-responsive phages also emerged as hubs, linking end of fast abundance shifts to network centrality. *Mushuvirus* was particularly notable: although its abundance declined at the end of the fast, its connectivity spiked at Day 10, then decreased at the 1-month follow-up, and increased again by the 3-month follow-up, at which point it became one of the highest-degree nodes, mirroring the network’s biphasic trajectory. While primarily infecting *Faecalibacterium*, it has also been linked to *Ruminococcaceae*. Moreover, *Mushuvirus* encodes a diversity-generating retroelement targeting a Hoc-like capsid protein, a feature that has been proposed to enhance phage adaptability within the digestive tract environment, potentially via enhancing phage binding to mucin in the intestinal mucus layer^92^. This could aid phage retention at the gut surface, increasing opportunity for interactions with bacteria^92^. In addition, these properties may allow *Mushuvirus* to influence also interactions at the mucosal surface^103^, potentially contributing to host defence by limiting pathogen adhesion and bacterial overgrowth^104^ and altering host-microbiome communication^105^. Thus, the emergence of *Mushuvirus* as a key network hub at the end of the fast and again by the 3-month follow-up, together with its consistent detection across cohorts, suggests a potential role in enhancing gut ecosystem resilience through the regulation of keystone bacteria such as *Faecalibacterium* and the reinforcement of mucosal defences.

Although overall diversity and composition appeared to return to baseline, the underlying network structure remained vastly different even 90 days later. This emphasises the importance of looking beyond diversity and abundance when evaluating microbiome change and its consequences for host health^106^. The GM functions as an ecosystem shaped by interactions that vary across individuals and time, and as such, stability, connectivity, and functional relationships are increasingly regarded as more meaningful indicators of health than taxonomic composition alone^107^. Therefore, this holistic exploration of the virome, phage-host dynamics, and cross-domain networks in response to fasting advances our understanding of how virome restructuring may intersect with host metabolic outcomes^108^, suggesting that fasting promotes a co-ordinated ecological reorganisation with potential implications for health.

### Strengths, limitations, and future directions

Key strengths of this study include the longitudinal, relatively large, and well-controlled cohort, and cross-cohort validation of viral responders. The use of Phanta and host linking also extends interpretability beyond viral taxonomic shifts. However, limitations should be noted. Firstly, the reliance on bulk metagenomics rather than VLP enrichment (unavoidable here given the retrospective use of the datasets) likely underrepresents low-abundance or free viral particles, whilst favouring the detection of viral sequences integrated within bacterial hosts^47,109^. Additionally, around 43% of detected taxa could not be assigned to known lineages, reflecting limits in current viral characterisation, potentially missing key players in viral dark matter. Moreover, methodological differences across cohorts, such as fasting protocols, refeeding strategies, sequencing depth, and laboratory workflows, might have introduced systematic bias, limiting direct comparison and validation of viral features across studies. Also, smaller participant numbers at follow-up limit statistical power. Finally, network inference was correlation-based at the genus level, so correlations may reflect shared niches rather than direct interactions. More extensive resampling with FDR correction and higher-resolution network analyses will be essential to uncover direct virus-bacterium relationships. Future work should also combine metagenomic and VLP sequencing, functionally annotate consensus phages or network nodes, integrate virome changes with clinical/metabolomic data to interrogate drivers of prophage induction, and whether viral shifts contribute to clinical outcomes.

## Conclusion

This study provides the first evidence that long-term fasting reshapes the human gut virome in structured and reproducible ways. Fasting induced a transient loss of viral diversity, a shift toward virulent lifestyles consistent with prophage induction, and specific changes in consensus phages.

Unlike inflammatory disease states, where *Faecalibacterium* phages expand, these patterns suggest nutrient withdrawal elicits a controlled and coordinated restructuring of microbiome dynamics, suggesting an adaptive rather than pathology-like trajectory. Fasting also promoted progressive reorganisation of virus-bacteria networks, with SCFA- and IPA-producing taxa and their phages emerging as recurrent hubs, and consensus phages such as *Mushuvirus* showing increasing connectivity and becoming among the most highly connected nodes by the 3-month follow-up. Together, these findings position fasting as a long-term controlled ecological reset, reorganising the virome-bacteriome interface, potentially contributing to host metabolic benefits. Although direct causality has not yet been established, integrating virome data with metabolomic and functional readouts will be key to testing these links. This work points to the potential of phage modulation, whether through fasting, diet, or targeted interventions, as a promising strategy to support metabolic and immune health.

## Additional Information

### Ethical approval

Metagenomic sequencing datasets and accompanying clinical data from two previously reported fasting cohorts at the Buchinger Wilhelmi Clinic (BWC), Überlingen, Germany, were re-analysed for this study: OralFast (NCT05449249) and GENESIS (NCT05031598). Ethical approvals were obtained for both studies from the State Medical Association of Baden-Württemberg. This ethics commission approved the study protocol of GENESIS on 26th July 2021. It was registered in an official clinical trial register on 2^nd^ September 2021 (ClinicalTrials.gov Identifier: NCT05031598). This ethics commission approved the study protocol of OralFast on 1^st^ of June 2022. It was registered in ClinicalTrials.gov on 4^th^ July 2022 (NCT05449249). Both studies were conducted in accordance with the Declaration of Helsinki and the guidelines of the International Conference of Harmonization of Good Clinical Practice Guidelines and German law.

## Authors’ contributions

NF was responsible for the design and execution of the analytical workflow, performed all bioinformatic and statistical analyses, created the figures, and wrote, edited, and structured the manuscript. RM conceptualised the project and co-supervised the project with AK. RM and FG designed and conducted the clinical study that generated the dataset used in this analysis. QD supported the project by advising on bioinformatics methods and data analysis. All authors reviewed, edited, and approved the final version of the manuscript.

## Data availability

Raw metagenomic sequencing data analysed in this study are available through the European Nucleotide Archive under the following accession numbers: PRJEB74035 (discovery cohort), PRJNA698459 (Maifeld et al.), and PRJEB75353 (Grundler et al.).

## Code availability

All relevant code to reproduce data analysis and visualisation will be made available on request.

## Consent for publication

The manuscript does not contain any personal data in any form.

This study was reported in accordance with The Strengthening The Organisation and Reporting of Microbiome Studies (STORMS) checklist^110^ (Table S8).

## Supplementary Tables

**Supplementary Table 1:** Metadata of all study participants.

**Supplementary Table 2:** Clinical marker changes before and after fasting.

**Supplementary Table 3:** Summary statistics and linear mixed-model comparisons for phage lifestyle metrics across timepoints.

**Supplementary Table 4:** Differential abundances of viral features (MaAsLin2 outputs) with sample prevalence.

**Supplementary Table 5:** Consensus fasting-associated viral species with cohort-specific effect sizes, significance values, and prevalence across all samples.

**Supplementary Table 6:** Viral species with predicted bacterial hosts and matched LMM responses to fasting.

**Supplementary Table 7:** Significant cross-domain virus-bacteria co-abundance correlations across timepoints (Spearman’s rho).

**Supplementary Table 8** - The Strengthening The Organisation and Reporting of Microbiome Studies (STORMS) checklist. (version 1.03; https://stormsmicrobiome.org)

## Supplementary Figures

**Figure S1:**
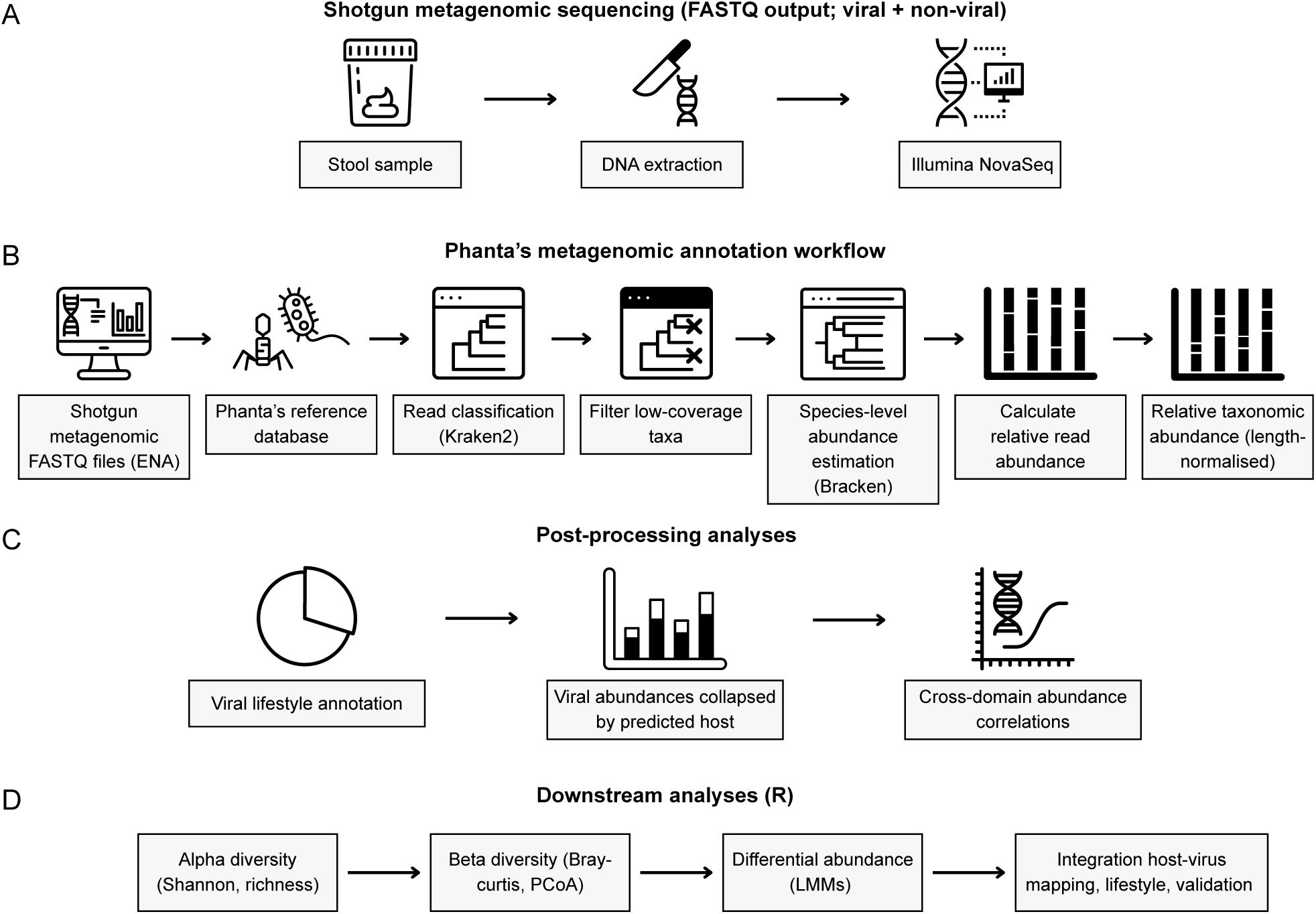
Overview of the metagenomic analysis from stool samples to statistical outputs. (A) Laboratory workflow: stool DNA extraction and shotgun metagenomic sequencing (Illumina NovaSeq) were performed previously as part of the original studies. (B) Bioinformatic workflow: FASTQ files were downloaded from the European Nucleotide Archive (ENA; study accession numbers provided) and processed with Phanta, which incorporates Kraken2-based classification, coverage-based filtering, and abundance estimation. All analyses were performed on the CLIMB cloud-computing platform. (C) Post-processing: Phanta-provided scripts were used to annotate viral lifestyle, collapse viral abundances by predicted host, and calculate cross-domain correlations. (D) Statistical downstream analyses: virus-filtered relative taxonomic abundance tables were analysed in R to calculate alpha and beta diversity across timepoints, perform differential abundance modelling using linear mixed models (LMMs). Integration included viral-host mapping, lifestyle statistics, and validation across other cohorts.

**Figure S2:**
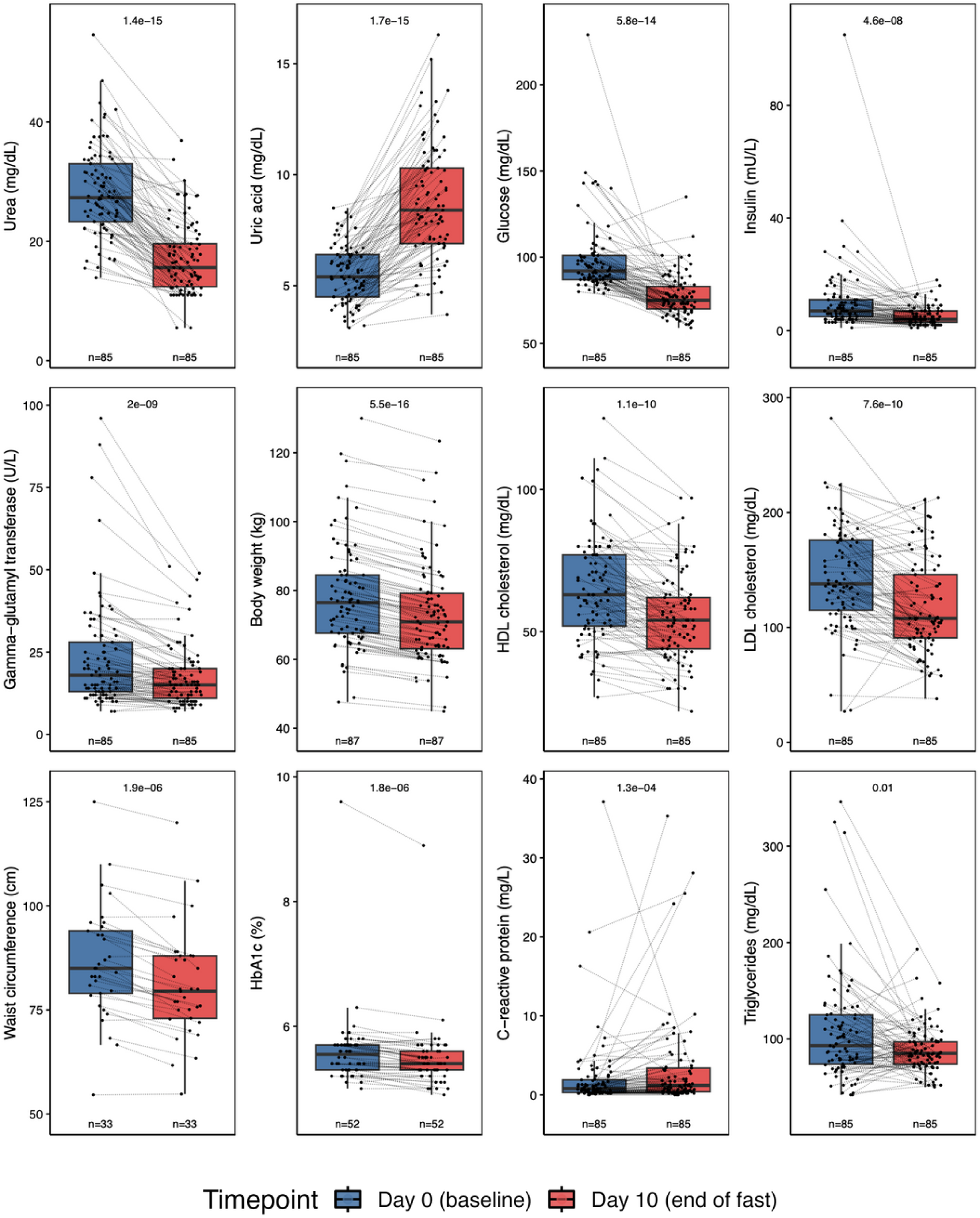
Changes in relevant clinical blood markers (reproduced and adapted from Ducarmon et al., 2024). Dotted lines connect the measurements within each participant from baseline to end of fast. P-values were obtained using a two-sided paired Wilcoxon test and were then underwent FDR correction with BH. Q-values are labelled on each plot. The boxplot centre value corresponds to the median, the box indicates the interquartile range, and whiskers extend to 1.5 times the interquartile range.

**Figure S3:**
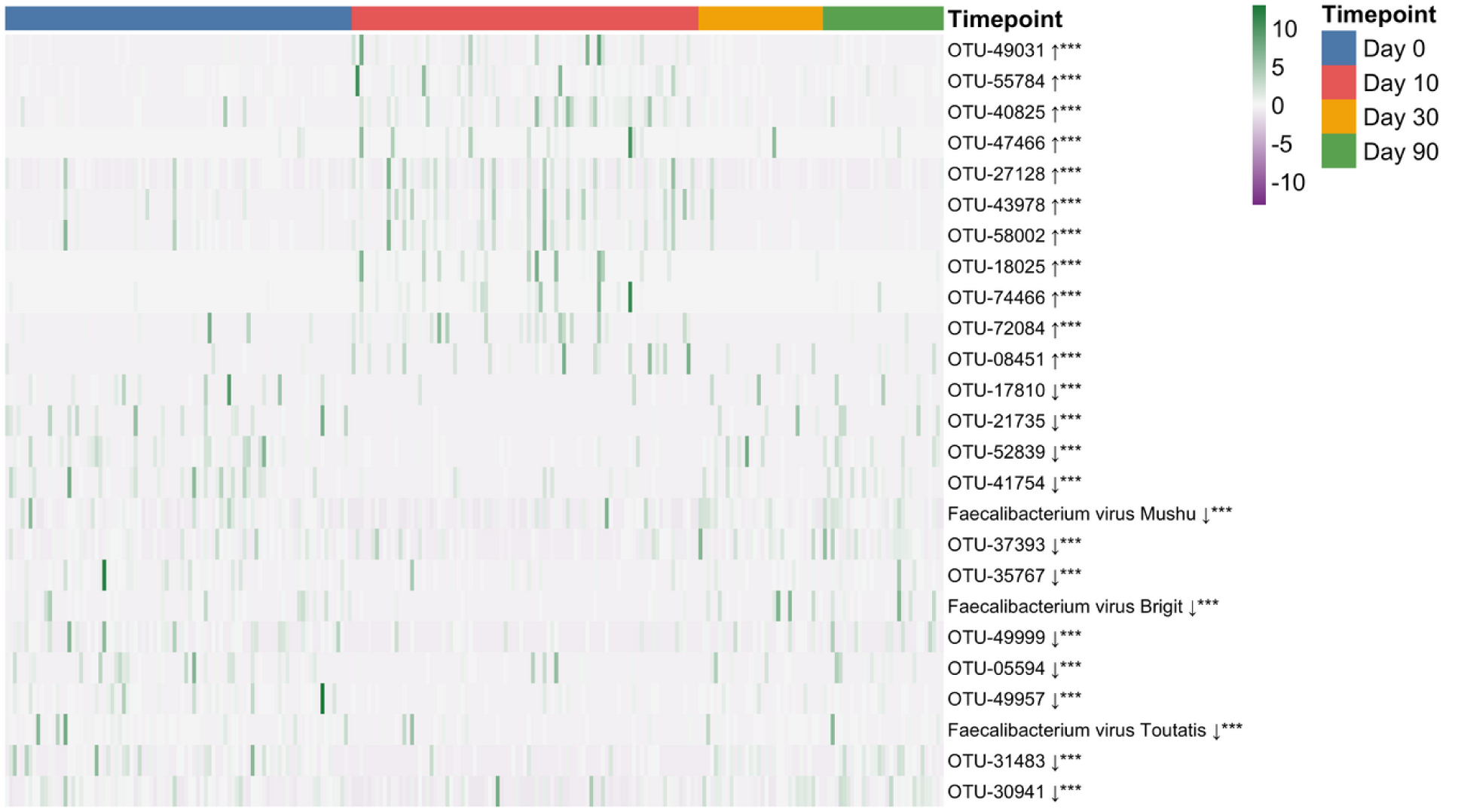
Heatmap of relative abundances for the top 25 fasting-responsive viral species across timepoints. Rows represent species (in the same order as A), columns represent individual samples ordered chronologically by timepoint. Abundances were row-scaled (z-score normalised) and visualised on a divergent colour gradient (purple = low, green = high). Arrows indicate the direction of change (↑ increase, ↓ decrease) based on model coefficients. Asterisks denote significance after false discovery rate correction (*p<0.05, **p<0.01, ***p<0.001).

**Figure S4:**
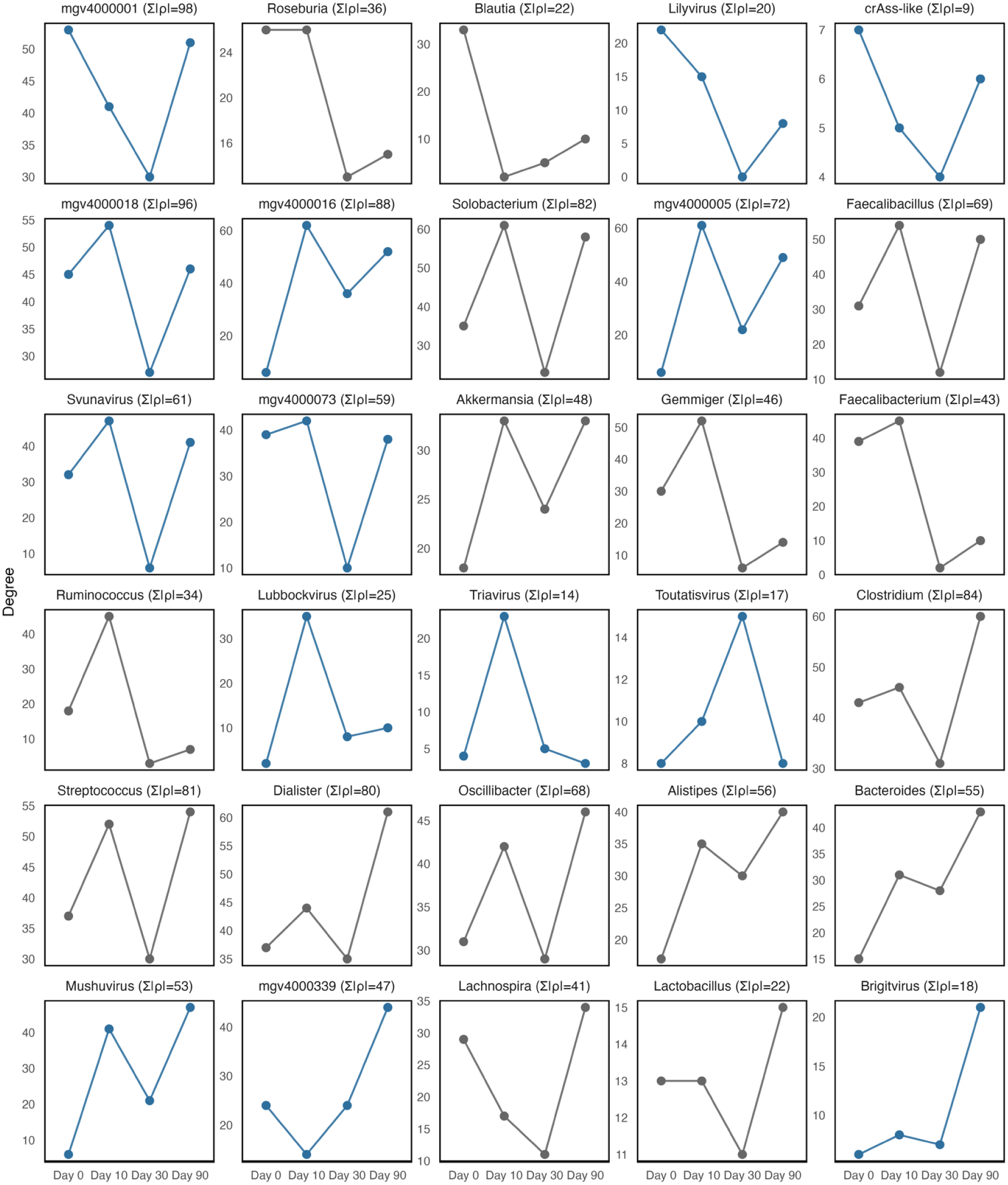
Temporal trajectories of selected viral and bacterial nodes in gut microbiome networks during fasting and follow-up. Line plots show node degree (number of virus-bacterium associations) across four timepoints: baseline (Day 0), end of fast (Day 10), one-month follow-up (Day 30), and three-month follow-up (Day 90). Viral nodes are shown in blue and bacterial nodes in grey. Strength values (Σ|ρ|) represent the cumulative sum of absolute correlation coefficients for each node, indicating overall interaction magnitude. Facets are ordered by the timepoint at which degree peaked, with stronger nodes listed higher within each timepoint group. Several crAss-like viral genera were collapsed into a single “crAss-like” node to represent this ecologically important clade.

